# Soft robotic steerable micro-catheter for the endovascular treatment of cerebral disorders

**DOI:** 10.1101/2021.06.28.21258767

**Authors:** Tilvawala Gopesh, Jessica H. Wen, David Santiago-Dieppa, Bernard Yan, J. Scott Pannell, Alexander Khalessi, Alexander Norbash, James Friend

**Affiliations:** Department of Mechanical and Aerospace Engineering, University of California San Diego, USA; Department of Neurosurgery, University of California San Diego, USA; Department of Radiology, University of California San Diego, USA; Melbourne Brain Centre, Royal Melbourne Hospital, Melbourne, VIC, Australia; Department of Surgery, University of California San Diego, USA

## Abstract

Catheters used for endovascular navigation in interventional procedures lack dexterity at the distal tip. Neurointerventionists, in particular, encounter challenges in up to 25% of aneurysm cases largely due to the inability to steer and navigate the tip of the micro-catheters through tortuous vasculature to access aneurysms. We overcome this problem with sub-millimeter diameter, hydraulically-actuated hyperelastic polymer devices at the distal tip of micro- catheters to enable active steerability. Controlled by hand, the devices offer complete 3D orientation of the tip. Using pressures up to 400 kPa (4 atm) we demonstrate guidewire-free navigation, access, and coil deployment *in vivo*, offering safety, ease of use, and design flexibility absent in other approaches to endovascular intervention. We demonstrate the ability of our device to navigate through vessels and to deliver embolization coils to the cerebral vessels in a live porcine model. This indicates the potential for microhydraulic soft robotics to solve difficult access and treatment problems in endovascular intervention.

## INTRODUCTION

Approximately one in fifty people in the US have an unruptured intracranial aneurysm, a thin-walled blister-like lesion on a cerebral artery that is prone to rupture. Studies estimate that cerebral aneurysms affect over 160 million people worldwide [93, 95], are increasing by over 5%/year [117], and are responsible for 500,000 deaths per year worldwide; half the victims are younger than fifty [11]. Annually, there are 30,000 new brain aneurysm ruptures in the U.S. alone [104]. Cerebral aneurysms and their consequent long-term medical complications impose an enormous direct economic burden of $500 million per year in the U.S.[12]. Intervention for unruptured aneurysms is generally known to be beneficial[122]; without treatment, over 50% of aneurysms larger than 5 mm eventually rupture and bleed[52, 53]. Of patients that suffer ruptured aneurysms, over half die [10, 45], and half the survivors experience long-term disabilities. The majority of aneurysms occur in vessels of 1.5 mm diameter or less [55].

Cerebral aneurysm treatment was limited to invasive surgical clipping until the clinical adoption of endovascular coil embolization in the 1990’s [37, 38, 36]. A minimally invasive approach, coil embolization involves inserting a micro-catheter at the femoral artery, navigating it through tortuous vessel anatomy under radiological guidance to the aneurysm via the aortic arch and carotid arteries, and deploying detachable coils into the aneurysm. The coils occlude the aneurysm from blood flow, inducing embolization. Endovascular coiling is now the most preferred[111], relatively cost-effective, and statistically more successful option for treating cerebral aneurysms[74, 73].

Catheters are the primary tool utilized for treating vascular pathologies such as cerebral aneurysms via endovascular approaches. Although catheter technology has improved over the past few decades, with notable advancements in *pushability* [9] and polymer coatings [77], the inability to steer the catheter tip *in vivo* remains [1]. This complicates the navigation of tortuous anatomy, gaining access into geometrically complex vascular pathologies that occur with aneurysms, and placement of the catheter tip in a stable position while deploying coils, stents or sophisticated implants. Current gold standard aneurysm embolization procedures use curved-tip guidewires to provide access via a previously introduced guidewire into aneurysm geometries that require acute catheter turns from the parent artery. However, upon retrieval of the guidewire in preparation to deliver coils, the catheter tip bends back to its original curvature, preventing coil deployment in the desired optimal direction.

The emergence of robotic-assisted surgical laparoscopic tools in the early 2000s—for example, the well-established, widely-accepted Da Vinci system [44, 103, 65]—demonstrates that robotic actuators can beneficially augment a surgeon’s dexterity in complex minimally invasive procedures. Likewise, the development of robotic tools for endoscopy and bronchoscopy have improved navigation and visualization of the gastrointestinal tract and pulmonary airways [90], and enabled advanced intervention for microscale suturing during such procedures [46]. Although now widely used in abdominal and thoracic surgeries that permit *>*1 mm endoluminal systems, steerable catheter systems have not yet been widely adopted in endovascular neuro-interventions.

Fabrication methods have advanced over the last half century to enable machining arbitrary shapes to the micro-scale. Progress in photolithography, nanoimprint lithography, twophoton lithography to electron-beam lithography and ion-beam machining have enabled the creation of massively parallel integrated semiconductor circuits, microelectromechanical (MEMs) devices, and micro and nanofluidics. These technologies have revolutionized medical devices and practices [67, 68]. However, manufacturing arbitrarily-shaped structures using hyperelastic materials at length scales between 10 *µ*m and 1 mm remains a challenge due to the convergence of van der Waals, electrostatic, gravitational, and other forces of similar magnitudes at this scale, making machining and manufacturing difficult [69]. Despite this, advancements in manufacturing methods have produced reductions in catheter diameters down to 1.8 Fr (600 *µ*m). This has enabled new endovascular techniques at more distal anatomical locations which is accompanied by increased tortuosity, additional bifurcations, and decreasing vessel diameters. This added complexity of distal vessel pathologies requires even more precise positioning for device deployment for effective treatment, requiring dexterous maneuvers by the interventionist [29, 123, 1]. As a result, robotic-actuator assistance has grown to become a pressing unmet need in interventional procedures.

Here we demonstrate how steerable endovascular microdevices can help address this clinical need, and how a combination of new and existing microfabrication techniques can be utilized to produce these devices. Due to their hyperelastic and deformable nature, soft-bodied hydraulic actuators offer a combination of safety and ability in traversing tortuous routes to a treatment location to then treat the patient, all by performing complex hand-controlled miniature movements in the vasculature that would otherwise be impossible or require an invasive route. Among endovascular procedures, a subset of specialist procedures requiring access of small distal vessels that would benefit from dexterous locomotion of submillimeter diameter catheters are illustrated in Figure 1.

**Figure 1:**
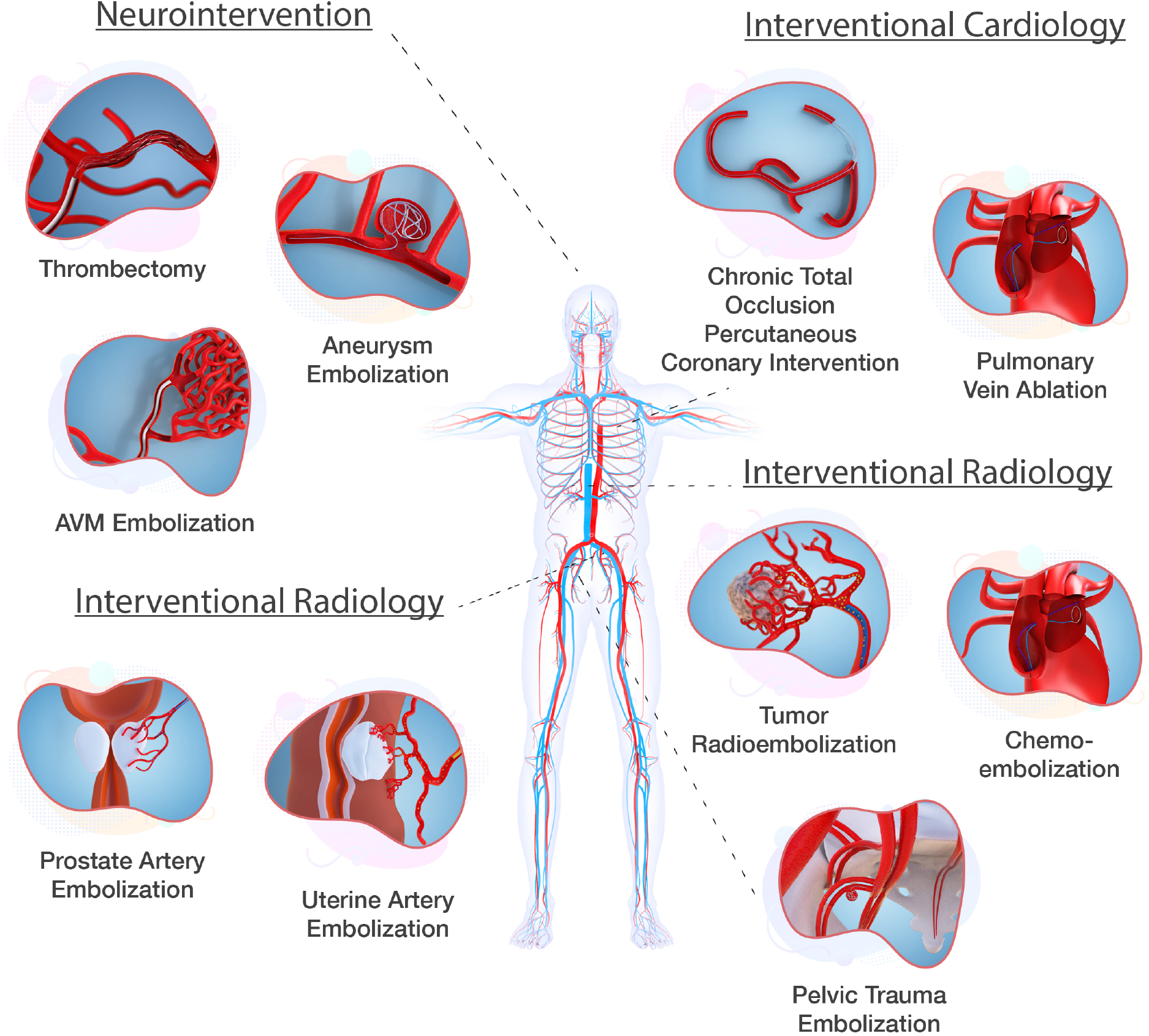
Endovascular procedures that would benefit from sub-millimeter diameter steerable catheters. Aneurysm embolization is the focus of this paper, but steerable sub-millimeter catheters are broadly useful across the human vasculature.

Endovascular coil embolization of brain aneurysms in neurosurgery is a key example, and our approach provides a means of controllably orienting a catheter in cerebral arteries to deploy coils in tortuous anatomical locations. Before we describe our approach and the results of using it, we review past attempts to serve this need.

Magnetic approaches to catheter steering first proposed in the 1950’s are constrained by magnetic field orientation, limitations of field saturation for small objects, and the complexity and cost of external field generation [105, 112]. Despite limitations [115, 106], studies on magnetically driven micro- and nanorobots have made valuable contributions towards steerable actuators [14, 18, 35, 15]. Magnetic actuation has been combined with soft materials to demonstrate micro-scale flagella [47], tunable microstructures [126], and externally controlled continuum soft robots [61]. Magnetically-driven steering has been demonstrated to be effective in cardiac electrophysiology procedures [22, 2, 27]. However, implementation requires costly infrastructure such as MRI magnets, robotic arms, and external control systems [94].

Pull-wire systems, based on the Bowden cable technique, were proposed as a means of catheter steering in the 1960’s [78, 41, 4, 107, 26]. Wires run from the proximal end of the catheter to the distal tip where a deflection can be achieved based on the push or pull motion of the cables. These range from single cable systems for unidirectional deflection [39] to multi-segment [118] for dual directional control. Pull-wire systems are hampered by inevitable undesired torsion [64], buckling [56], internal friction [9], and external friction [83, 13]. These factors, and the stiffening often used to avoid them, can independently or together result in undesired tip motion resulting in vascular damage [7, 32, 120].

Piezoelectric ultrasonic approaches [124, 17] have also been proposed, but currently face practical reliability and safety challenges. Using vibration transmitted across a contact interface to a steerable component, the devices are liable to lose the steerable component or exhibit unanticipated motions when in contact with the vascular wall. They also require 1 MHz or greater frequency oscillating electrical signals to be transmitted to the distal tip; neither wired nor wireless methods are able to do this to date.

The burgeoning field of soft robotic methods drawing deformable materials into bioin-spired 3D structures has seen growth in research and applied technology [21, 98, 66, 92, 121] ranging from anatomical replacements such as prosthetic limbs to surgical tools [24, 116, 19, 5, 30]. Studies have demonstrated the use of soft robotics in balloon catheters with electrophysiological and ablation capabilities [60], and in implantable soft robotic sleeves to support heart function [97]. Soft robotic based hydraulic and pneumatic actuators first proposed in the 1990’s [108, 109, 110] include pressure-driven motion [50], vacuum-driven motion [40], and hydraulic jets [31]. Through newer fabrication capabilities, smaller soft robotic structures with unidirectional motion [85] and more complex larger scale soft robotic structures with untethered actuation capabilities [119] have been presented.

There are over eight hundred patents that describe methods of enabling steering at the distal end of a catheter tip [29, 1]. A small number of these that utilize magnetic [86, 3] and pull-wire systems [99] are commercially available, but at diameters greater than 1 mm. There has to date been no demonstration of a steerable sub-millimeter diameter, soft-bodied micro-catheter for endovascular procedures. For clinical use, a micro-catheter must be radioopaque to be visible during a procedure and have a central lumen with a lubricious coating to permit the smooth passage of guidewires and implants. The ability to actively steer, navigate, position and deploy sophisticated implants such as coils, flow diverters, stents, or glue does not exist at these scales today.

Here we present a soft polymer tip distally attached to a 1.6-m catheter with a contiguous lumen. Since balloon catheters—representing a crude form of hydraulic soft robotics—are broadly accepted as safe and effective in endovascular intervention, a hydraulically-actuated fully steerable micro-catheter can reasonably be expected to be similarly safe and effective if the pressures used are similar or less. The tip in our device is hydraulically steered via four 50 *µ*m-diameter cylindrical, saline-filled channels, in part inspired by highly deformable micro-scale mechanisms observed in nature [71]. These axially-oriented channels are equally spaced around the catheter wall for hydraulic actuation. Pressurizing one or more of these channels produces an internal stress which creates an axial differential strain and an associated deflection, and thus “steers” the tip away from the pressurized channel(s) in a desired direction [28]. While there is also radial and azimuthal internal stress from the pressure, the structure is specifically designed to minimize radial expansion and promote axial deformation, notably unlike balloon catheters. This prevents inward expansion to potentially constrict and clamp whatever may be in the internal lumen, or outward expansion to potentially occlude the vessel or damage the arterial endothelium.

The steerable tip was fabricated using a simple but effective lab-based technique to manufacture sub-millimeter scale high aspect ratio structures with hyperelastic materials (detailed in Materials and Methods). Both the inner lumen and outer surface of the catheter were coated with a hydrophilic coating to reduce friction between the micro-catheter and the endothelial walls of the arteries, and between the micro-catheter and devices such as coils passed within the lumen. The tip was assembled with 160 cm-long catheter-grade tubing connected to a handheld controller to form a closed-system, stand-alone micro-catheter with a steerable tip that can be used independent of external controllers, robotic arms, or additional infrastructure.

We demonstrate the use of the active steering capability of this micro-catheter device in navigating through tortuous cerebral vasculature and in deploying coils to treat cerebral aneurysms in a representative *ex vivo* model. The results and interventionist feedback from testing in the *ex vivo* model were used to iteratively improve the device design to eventually produce a device suitable for *in vivo* testing. A representative *in vivo* endovascular coiling procedure was conducted in live porcine. Contrary to conventional wisdom that tethered micro-actuation at the high length-to-diameter aspect ratios required in medical applications—10000:1 in our case—would be impractical due to viscous losses, we demon-strate that well-designed structures with engineered hyperelastic materials can enable seamless real-time control of micro-scale soft bodied robots via 1.5-m long, 50 *µ*m diameter microfluidic channels.

## RESULTS

### A hydraulically actuated soft robotic steerable tip at dimensions compatible with cerebral arteries

As previously noted, a majority of cerebral aneurysms occur in vessels less than 1.5 mm in diameter [55]. Cerebral arteries supplying blood to the brain begin above the neck, at the base of the internal carotid artery (ICA) through to the middle cerebral artery (MCA) bifurcation, next branching to smaller and more tortuous cerebral vessels. The MCA has a diameter of approximately 2.6 mm [100] (Fig. 2 a); blood vessel branches beyond the MCA gradually decrease in size to diameters of 1 mm or less [48]. To navigate smoothly within cerebral vessels, current gold standard micro-catheters have an outer diameter of 900 *µ*m (3 Fr) and inner diameter of 400 *µ*m (1.2 Fr) to provide a lumen for guidewires, detachable coils, and sophisticated endovascular devices such as stents and flow diverters. To provide clinical compatibility and immediate familiarity when used in existing procedures, the steerable tip micro-catheter described here was designed to have an outer diameter of 900 *µ*m, enclosing four 50 *µ*m diameter channels for hydraulic actuation and a 400 *µ*m diameter central lumen. Upon inflating one of the 50 *µ*m diameter channels, an axial differential strain is induced in the structure about its mid-plane—the axis of symmetry along the length of the tip—causing bending of the tip away from the inflated channel (Fig. 2 b). The fabrication of the steerable tip is illustrated in Fig. 2; details are provided in the Materials and Methods. A meso-scale molding process employing thin wires cast with silicone rubber in polyurethane plastic molds (Fig. 2 c) was used.

**Figure 2:**
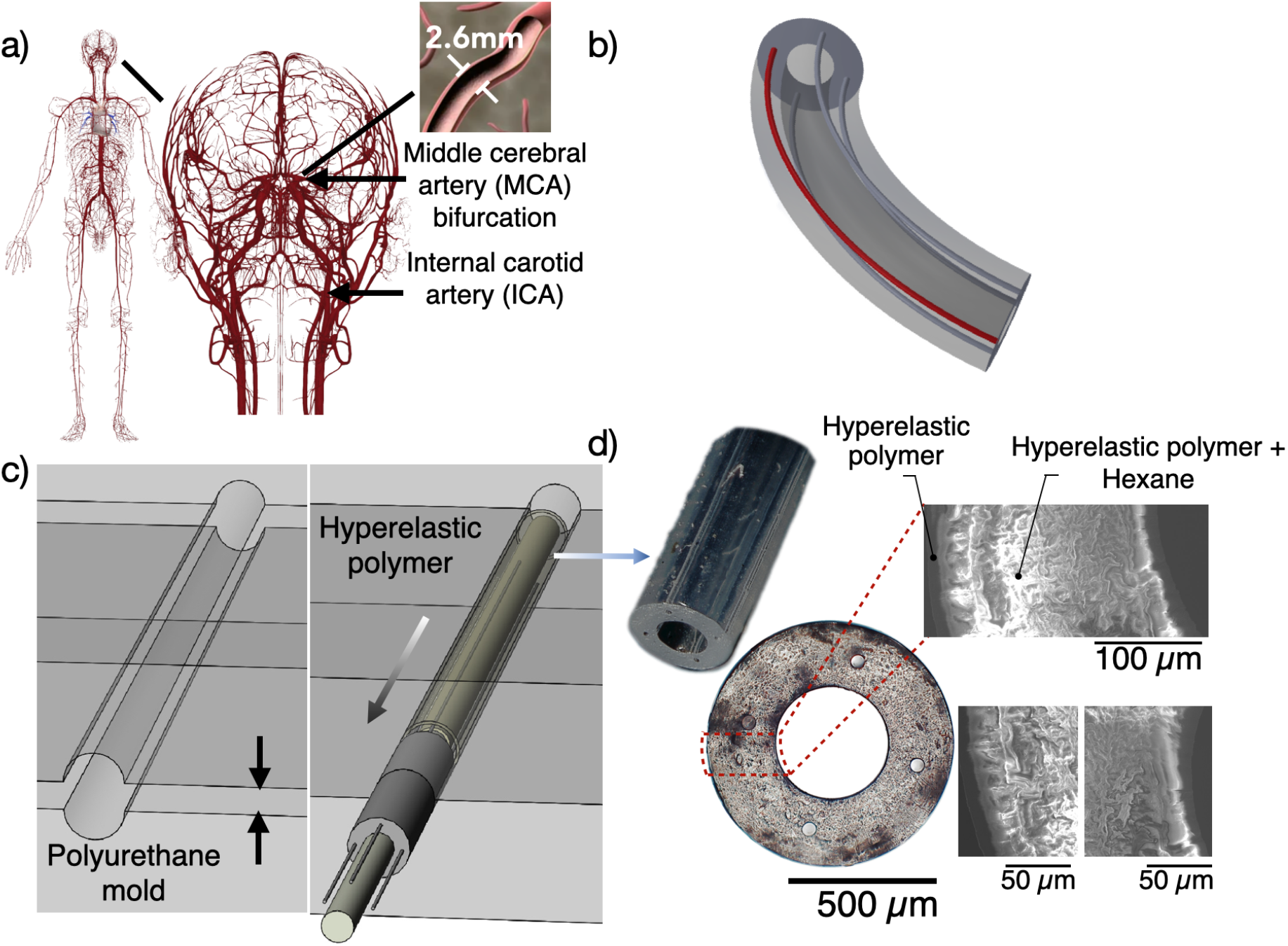
Engineering a steerable catheter. a) Illustration of the cerebral arterial structures and the average diameter of the middle cerebral artery. b) Design and working principle of the hydraulically driven tip: when a channel is inflated (indicated in red), the tip deflects in the opposite direction. c) Fabrication of the catheter is a multi-step molding process.d) Plan and cross-section images of the as-fabricated steerable tip tubing, with a length of 15mm, an outer diameter of 900 *µ*m, an inner diameter of 400 *µ*m, and four 50 *µ*m channels in the tube wall. The relatively rigid coating upon the softer interior material that forms the hydraulically-driven tip is visible via SEM cross-section images, and is approximately 25 *µ*m thick.

### The steerable tip is engineered to limit radial expansion and promote bending

Unlike the use of saline injection into catheter tips to radially inflate balloons to pin a catheter and deploy and expand a stent, avoiding radial expansion of the fluid-filed channels is important in our device. This was achieved by carefully designing the placement and size of the fluid channels, and by co-casting concentric layers of a platinum-cure hyperelastic silicone rubber (Dragon-Skin^®^ 10 SLOW, Smooth-On, Inc., Macungie, PA USA; details provided in Materials and Methods).

### Choosing the radius and radial location of the hydraulic channels

While the inner and outer diameters of the steerable micro-catheter tip are inherently constrained by the diameter of the cerebral arteries [100] and designed to be commensurate with the dimensions of the existing gold standard micro-catheters, producing a steerable tip without radial expansion required a design study focusing upon the radius, *R* and radial position, *R*_P_, of the hydraulic channels as parameters. Using a computational model, the radii for hydraulic input were varied from 25 *µ*m to 100 *µ*m while maintaining a fixed radial position (325 *µ*m from the center). The position of the hydraulic channel was varied from 260 *µ*m to 390 *µ*m from the center of the lumen, while maintaining a fixed channel radius of 50 *µ*m. The number of channels also impacts the device mechanics, as a greater number of small channels could produce an improved ability to steer, but the significant drawback in introducing more than three to four channels is the rapid increase in complexity of the tip-to-catheter and catheter-to-hand controller interfaces. Consequently, we limited our study to four channels, and noted that three or fewer channels produce similar results, albeit with reduced functionality with two or fewer channels. Furthermore, the computational expense of accurately computing the nonlinear finite deformation of a non-linearly (stiffening) elastic media in a structure with a length:width aspect ratio of 1:1000, together with practical fabrication limitations imposed by the small scale and intrinsic challenges in making such structures, limited our choices for the channel radius, *R*, and the radial position of the channel, *R*_P_ to three values each. Even so, it is apparent from Fig. 3b,c (and Fig. 4) that the range of choices considered have a significant effect on the performance of the device.

**Figure 3:**
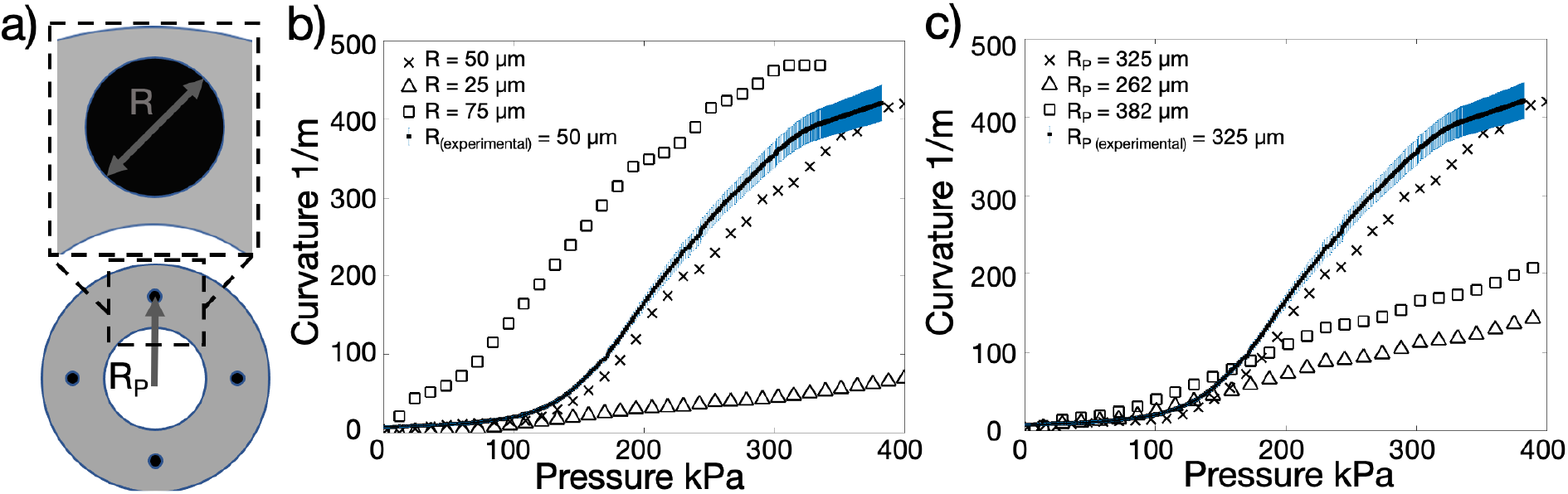
Design parameters of the steerable tip, and the consequent curvature as a function of input pressure. Nonlinear finite deformable structural mechanics analysis was used to compute the relationship between the tip curvature and input pressure for three values of a) the channel radius, *R*, and the channels’ radial position, *R*_P_. The b,c) curvature of the tip as a function of the input pressure produces significantly different computational results for the b) radius *R* with the radial position *R*_P_ at *R*_P_ *=* 325 *µ*m, and c) radial channel position *R*_P_ for a channel radius *R =* 50 *µ*m. The computed results are similar to (solid line in b,c) experimental results obtained with *R =* 50 *µ*m and *R*_P_ *=* 325 *µ*m. The experimental results are averaged for four channels, blue lines indicate error bars (standard deviation).

**Figure 4:**
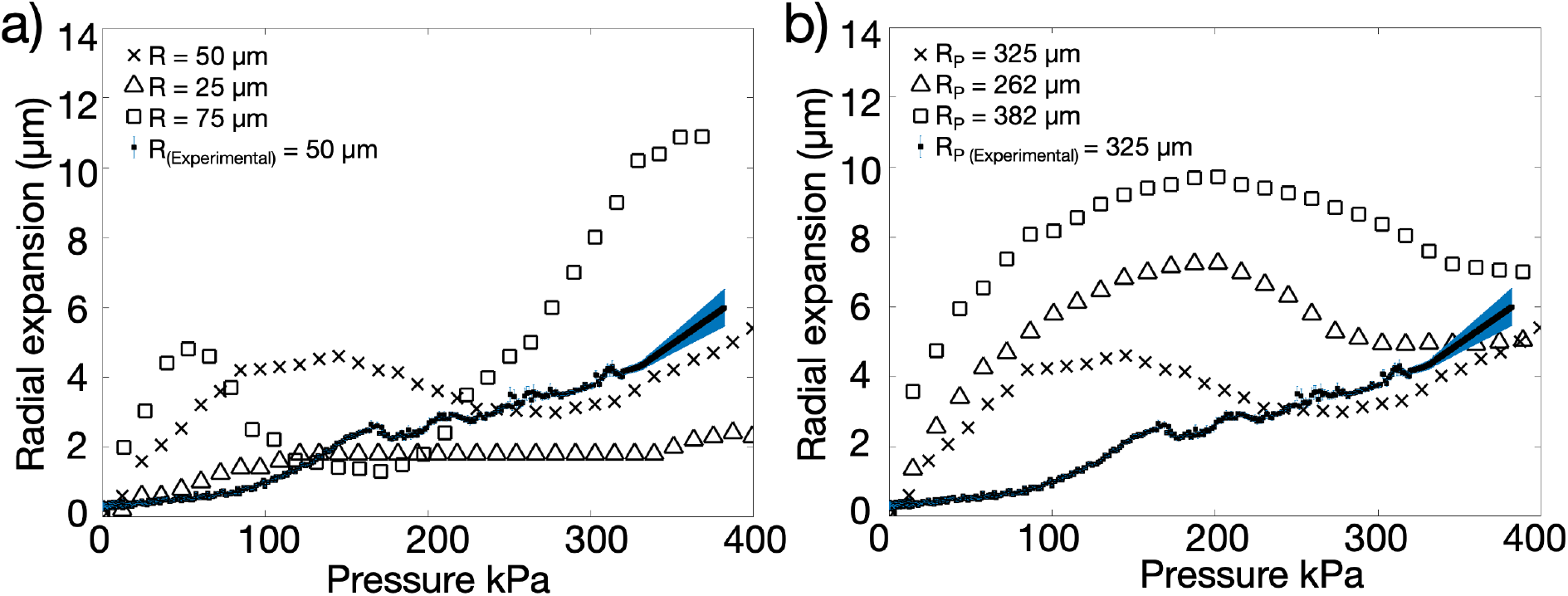
The hydraulic channels’ position and radius affects the relationship between the tip’s radial expansion and the input pressure. Computed radial expansion of the steerable tip at its outer surface as a function of a) channel radius *R* with *R*_P_ *=* 325 *µ*m, and b) channel radial position *R*_P_ with channel radius *R =* 50 *µ*m, compared to experimental results for a tip with *R =* 50 *µ*m and *R*_P_ *=* 325 *µ*m. The experimental results are averaged for four channels, blue lines indicate error bars (standard deviation).

**Figure 5:**
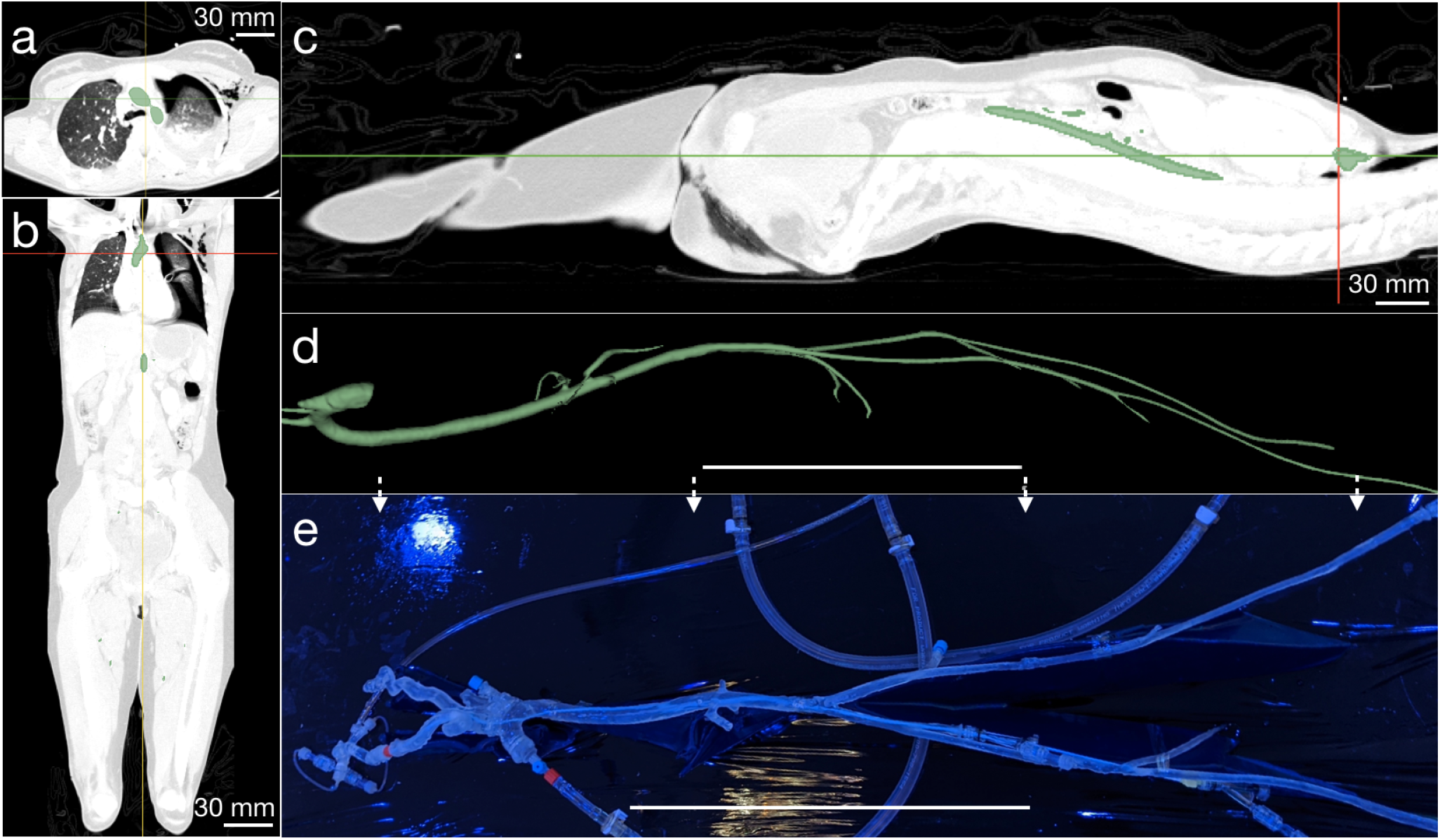
Fabrication of an *ex vivo* silicone model from anonymized patient data. a) Axial, b) sagittal, and c) coronal views of CT angiogram images with vascular segmentation for a single slice shown in green. This process produces a d) complete vascular model from the femoral artery to the aortic arch, including principal vascular branches, in particular the carotid arteries to the cerebrovasculature. Through 3D printing and lost ABS casting as detailed in the text, e) a functional, fluid-tight vascular model may be produced. (d,e) Scale bar = 25 cm.

Further, Fig. 3b,c: solid lines) plots the tip’s curvature versus input pressure using image capture and post-processing. The pressure required to achieve a curvature of 400 1/m (180*^◦^* total bend, forming a “C” shape) is 350 kPa, significantly lower than traditional balloon catheters (600 kPa) [43] for stent deployment. Positioning the radius away from the midpoint of the wall thickness results in inadequate curvature at higher input pressures (Figure 3 b). The 50 *µ*m diameter channels exhibit 10% strain to achieve a 180*^◦^* bend. The materials used have a strain at failure *>*350% [34] at pressures 4X that required to achieve 10% strain. In the inadvertent case of over-pressurizing the hydraulic channel, the steerable tip loops on itself at pressures higher than 350 kPa, and will rupture at a pressure of 1200 kPa thus providing a safety factor *≈*4.

While a smaller channel radius of *R =* 25 *µ*m, for example, helps to reduce radial expansion (Fig. 4 a), the bending curvature is significantly reduced for the same input pressure (Fig. 3 a). A larger radius, *R =* 75 *µ*m, produces greater curvatures at relatively lower input pressures (Fig. 3 a), although radial expansion is also significantly greater (Fig. 3 a). The large mismatch in the simulated results for the radial expansion indicates that, initially, the hydraulic channels rapidly expand and reach a maximum saturation point where the concentric layers prevent further expansion. As the hydraulic channels elongate, the radial expansion reduces to compensate. Although the experimental results demonstrate a similar trend, it is more subtle than the simulated results. While the concentric layers prevent excess radial expansion at the required curvatures, the elongation during the experiment does not cause a reduction in the hydraulic channel radius.

### Tailoring the tip’s construction to further reduce radial expansion

The innermost and outermost surfaces are made with the hyperelastic polymer, while the region enclosed by these surfaces is formed from a relatively softer 1:1 mixture by weight of the hyperelastic polymer with hexane (CAS 110–54–3, 95% anhydrous hexane, SigmaAldrich, St. Louis, MO USA).

The purpose of this layering and the placement and sizing of the fluid channels is to promote axial deformation while minimizing radial expansion. The key is to observe that the layering of different hyperelastic materials induces stress-stiffening [34]. Consequently, the introduction of a pressure in one of the channels produces radial, axial, and azimuthal stress, and as these values increase, the rate of strain increase gradually falls. The radial strain, over the relatively short length scale of the wall thickness of the catheter tip, quickly reaches the stiffening portion of the hyperelastic stress-strain response.

However, because the tip is *∼*100 times longer than its radius, the axial deformation is likewise *∼*100 times greater than the radial deformation for the same strain value. The axial deformation is therefore far greater, producing ample bending of the tip with negligible radial expansion. This material choice is contrary to the choice of strain-softening hyperelastic media typical in balloons. Further, the relative rigidity of the polymer without hexane treatment serves as a constraint to radial expansion, squeezing the relatively soft hexane-treated polymer within to axially exude from the channel pressure. Catheter tips made without layering exhibit significantly larger radial expansion (Fig. 8).

### Forming full-length (160 cm) catheters

The soft polymer tip with a 400 *µ*m diameter central lumen and 50 *µ*m diameter hydraulic channels at a radial position *R*_P_ *=* 325 *µ*m was attached end-to-end to long catheter-grade and matched multibore tubing (as detailed in the Materials and Methods) to produce 160-cm long catheters that mimic current micro-catheters used in clinical treatment of cerebral aneurysms. Pebax, a copolymer made of polyamide and soft polyether used in the majority of commercially available catheters, was used in the multibore tubing with graduated stiffness along its length. For the larger and stiffer femoral artery and aorta, high and medium durometer Pebax was used for the proximal catheter, while lower durometer Pebax was used for the distal catheter passing through the smaller and more fragile cerebral arteries.

### Biplane x-ray visible radiopacity

To ensure the steerable tip is visible via digital biplane x-ray typical in interventional angiography suites, gold radiopaque markers (27121A, Johnson Matthey, San Diego, CA USA) were embedded in the soft polymer tip. Markers are positioned at three distinct locations. The first and second markers indicate the beginning and end of the steerable portion of the micro-catheter tip. The third marker, the most proximal, is located 3 cm from the distal end of the catheter tip and aligns with the marker on the coil pusher of an embolization coil. This third marker aids in visualization of the point at which an entire coil has been deployed into an aneurysm, indicating that the pusher should not be advanced further. This marker could be similarly used for other types of devices, including flow diverters or stents that may be placed in the lumen of the micro-catheter. The remainder of the non-steerable portion of the catheter was made with standard barium-filled Pebax tubing to achieve clinically required radiopacity.

### Reducing friction between the inner lumen and guidewires and coils

To overcome friction between the catheter’s inner lumen and devices such as stents or coils as well as friction between catheters and the surrounding vessel, commercial catheters have hydrophilic coatings on the inner lumen to provide a lubricious passageway for devices, and on the exterior to reduce the risk of damage to the vascular endothelium and to avoid vasospasm [88]. The exterior coating furthermore reduces sudden release of built up forward pressure accumulated in frictional contact between the catheter and vascular wall at multiple vascular bends. As catheters are advanced, frictional resistance on the outer catheter surface at these bends may clamp it and cause it to accumulate elastic energy that can be suddenly and unpredictably released to pop the distal portion of the catheter forward, potentially resulting in catastrophic vessel dissection or aneurysm rupture. Silicone rubbers similar to those used in this study are known to exhibit significant surface friction [89]. To reduce this friction we employed a hydrophilic coating (ON–470, Aculon Inc, San Diego, CA USA) to achieve smooth advancement and navigation of our micro-catheter through both *ex vivo* and *in vivo* models, and the smooth advancement of coils through the micro-catheter lumen.

### An *ex vivo* model to provide practice and feedback in treating cerebral aneurysms with this system

Performance of the steerable catheter was assessed and iteratively improved through testing by clinical practitioners in *ex vivo* silicone models representative of the human vasculature, similar to models used to test commercially available catheters [72, 80, 63]. A single vasculature model of vessels from the femoral artery to the internal carotid artery (ICA), including the aortic arch, was constructed. Anonymized CT-angiogram DICOM images from a patient with representative vessel geometry were segmented, 3D printed, and used to fabricate a representative model using silicone rubber (details provided in Materials and Methods). A separate silicone phantom from the internal carotid to the cerebral vessels including the aneurysm was fabricated using the same approach; the aneurysm presented here was selected to represent a challenging geometry at the posterior communicating artery (PCOM) requiring acute angle turns. The primary factors iteratively improved include transitional stiffness, friction and radio-opacity.

### Neuroendovascular surgical setup commensurate with clinical procedures

Using the right femoral artery of the silicone model as the access point, a guide catheter was inserted into the silicone model; the distal end of the larger guide catheter was parked at the mid internal carotid artery (ICA). Standard saline was used to flush the guide catheter and micro-catheter in line with standard endovascular neurosurgical protocols [57]. Unlike conventional endovascular neurosurgery, once the guidewire was used to reach the aneurysm location in tandem with the micro-catheter, the guidewire was retrieved and hydraulic actuation of the steerable catheter engaged to direct the distal end of the steerable tip into the dome of the aneurysm then was used to lock the tip at the geometric center of the aneurysm. Detachable coils were introduced through the hub of the steerable micro-catheter and deployed into the aneurysm dome (supplementary video 2).

### Testing *in vivo* in porcine

This study was approved by the University of California San Diego Animal Care and Use Committee. A skeletally mature (47 kg) female pig underwent general endotracheal anesthesia. Cardiorespiratory monitoring was performed throughout the procedure under the supervision of a veterinary technician.

Referring to Fig. 6, after surgical exposure, a 6-F sheath was placed in the left femoral artery using the modified Seldinger technique [102]. Under road-map fluoroscopic guidance, a 5-F guide catheter was advanced to the right common carotid artery (CCA) over a 0.035 in guidewire. The 0.035 in guidewire was retrieved. A straight 0.014 in micro wire was inserted into the soft-robotic steerable catheter and the microsystem was then advanced to the distal end of the guide catheter. The 0.014 in micro wire was retrieved and hydraulic actuation of the steerable catheter was engaged to traverse from the external carotid artery to the ascending pharyngeal artery. Hydraulic actuation was disengaged, returning the tip to its native, straight orientation. The steerable micro-catheter was further advanced and hydraulic actuation, applied by pressurizing saline through a 1 ml syringe was then engaged to direct the distal end into the parotid artery. The steerable tip was locked in a fixed curvature position using a one-way stopcock and detachable coils were deployed via the hub of the micro-catheter (*see* supplementary videos 3 and 4).

**Figure 6:**
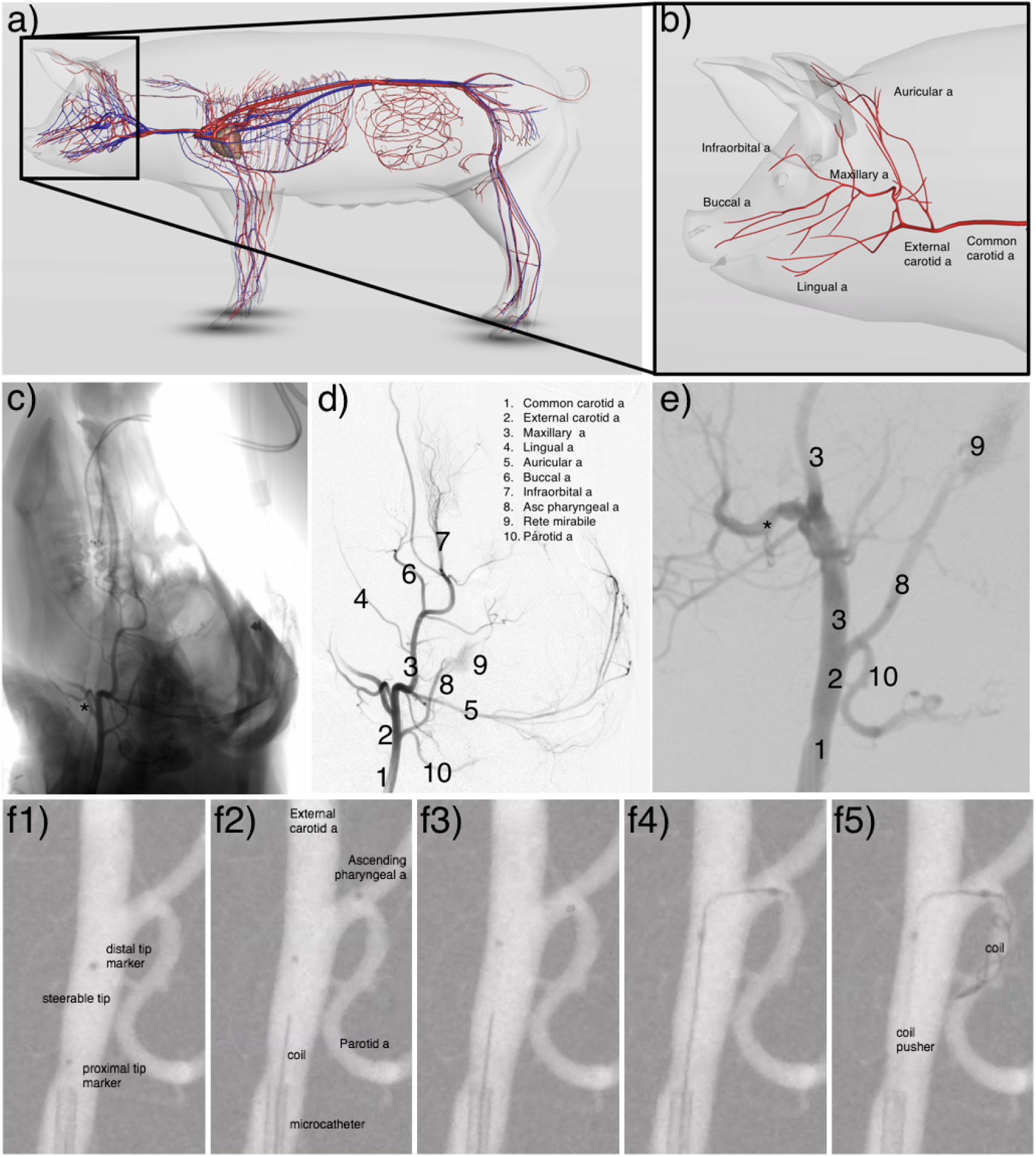
Animal study. Illustration of porcine (3D Pig anatomy illustration by Biosphera) a) arteries and b) cerebral arteries for reference. (c) Anteroposterior (AP) diagnostic subtraction angiogram of the right CCA. (d) Spot AP road-map fluoroscopic image of the right CCA. enlarged in (e). (f1) Position one, showing the steerable microcatheter prior to introduction of a hydraulic load. (f2) Position two, showing the micro-catheter after hydraulically steering the tip to access the ascending pharyngeal artery from the external carotid artery. (f3-4) Position three, showing the hydraulic steering of the catheter tip to access the parotid artery from the ascending pharyngeal artery. (f5) Coils deployed in the parotid artery from the stabilized catheter tip.

## DISCUSSION

Here we introduce the design and fabrication of a hydraulically actuated soft robotic microcatheter, and demonstrate its ability to steer in a pig model and enable deployment of coils in complex anatomical geometries with acute angles.

Although over 50% of intracranial aneurysms are treated via endovascular coiling, the efficacy and outcomes of endovascular aneurysm embolization are still inferior to traditional invasive clipping [23]. The difficulty in navigating a micro-catheter through tortuous vasculature, directing the catheter tip to the aneurysm dome, and holding it in a stable position account for many coil embolization procedure failures. These include incomplete aneurysm occlusion [101], aneurysm recanalization [91], coil malposition [58], and intraoperative rupture [25, 62]. These failures can be attributed to unfavorable vessel tortuosity and aneurysm geometry. Furthermore, up to 25% of intracranial aneurysms cannot be endovascularly treated, often due to aneurysm location — where the vessel is too difficult to reach, or geometry — where the dome, neck, or angles are unfavorable for micro-catheter cannulation or coil support. Up to 25% of surgical neurointerventions fail in attempted endovascular treatment of intracranial aneurysms [23, 101]. The ability to treat aneurysms of unfavorable shape and size, and the durability of embolization coils in the dome post intervention remain challenges in the endovascular coiling technique [91].

In cases of severe carotid system tortuosity, intracranial positioning of micro-catheters to access the aneurysm dome may be impossible or hazardous. In tortuous or fragile vasculature, turns of 180 degrees and 360 degrees are particularly difficult, catheterization often requires multiple attempts, and vasospasms are often induced [88]. Tortuous arterial anatomy forces micro-catheters along the outer curvature of the vessels at each turn, creating stresses on the vessel, a potential mechanism for vessel dissection [8]. Furthermore, minimizing overall procedure time is important to not only minimize costs, but to avoid excessive x-ray fluoroscopy exposure and associated risks such as thromboses or tumor growth [84, 54, 87].

To address the lack of steerability, catheter manufacturers provide 45°, 90°, and C-shaped tips in fixed orientations, and neurointerventionists will often retrieve, reshape, and reintroduce their microguidewire tips in hopes of accessing tortuous aneurysm geometries using a “trial and error” approach. Upon retrieval of the guidewires, the catheter tip will return to its native shape and none of the catheters can be steered once inside blood vessels.

A micro-catheter with a soft tip that can be controllably directed in an artery during endovascular coil embolization could decrease procedure time and reduce the failure rate of these procedures. Comparative studies between steerable and non-steerable catheters have demonstrated significant reductions in procedure time and improvements in patient safety [114, 42, 70]. Steerable catheters are known to enable better positioning of sophisticated implants [82, 113] and successful treatment in patients who would otherwise be precluded from endovascular treatment with a standard non-steerable catheter [51, 94].

The work described here demonstrates the use and efficacy of a hydraulically actuated soft polymer tip that can be steered and locked in a desired position at the distal end of a micro-catheter *in vivo*. A micro-catheter with a steerable tip would allow safer and quicker navigation through tortuous anatomy, improved access of a more desirable target location relative to the aneurysm dome, and coil deployment with a more stable platform. Due to their inherent rigidity and size, traditional robotic tools would not be able to achieve this within blood vessels at the millimeter scale. Likewise, a purely soft material approach would present serious technical challenges with regard to pushability *in vivo*. Here we leverage microfabricated soft robotic actuators with engineered hyperelastic materials to assemble full length catheters with transitionally variable stiffness from the distal to proximal end. This device further enables the deployment of devices such as stents and flow diversion pipelines in challenging anatomical configurations. Although balloon assisted coiling [76] has been used to mechanically assist in orienting a catheter tip in endovascular neurointervention procedures, this method does not provide control over catheter tip orientation within the aneurysm dome, and due to anatomical size constraints, cannot be used in smaller blood vessels.

In the present work, a steerable tip micro-catheter at the submillimeter length scales commensurate with endovascular neurosurgical catheters was fabricated, characterized, and tested in *ex vivo* silicone models representative of the human vasculature. Feedback from practicing neurointerventionists was incorporated to iteratively improve device performance before conducting an *in vivo* trial in live porcine. Existing technology is unable to systematically provide access to arteries that are acutely joined to the parent artery where the catheter is located. Here, we have shown the ability to accomplish this difficult task with the steerable tip *in vivo* through the following steps (referring to Fig. 6 and supplementary movie S2):

1. Access the ascending pharyngeal artery from the external carotid artery,
2. Access the parotid artery from the ascending pharyngeal artery,
3. Maintain the acute turn position while deploying coils.

Furthermore, the new steerable catheter demonstrated a higher level of safety than clinically experienced when using current gold standard catheters. A significant risk of endovascular platforms is mechanical injury to the vessel. This manifests as a vessel dissection and/or perforation which is immediately evident to the surgeon during follow-up angiograms performed during the procedure. Our catheter caused no injury to the endothelium or perforation of the vessel lumen on follow up angiographic views as shown in Figure 6 (f1-f5, supplementary movie S2). Based on these three characteristics, there is a reasonable basis to conclude that our catheter may provide a greater level of procedural safety relative to current gold standard procedures. However, this still has to be verified through a statistically significant number of animal trials outside the scope of the present research effort.

To our knowledge this was the first in vivo demonstration of a guidewire-free navigation, access, and coil deployment procedure in cerebral blood vessels using a hydraulically actuated micro-catheter. The main purpose of the in vivo model was to demonstrate our catheters’ performance in physiologic conditions including temperature, blood pressure/pulse pressure variation, and the complex chemical composition (and prothrombotic characteristics) of live blood. This proof-of-concept experiment demonstrated that our catheter performed as intended.

Based on the resulting data, the technology demonstrates promise as a viable approach to augment the function of existing micro-catheters in treatment of vascular injuries. Likewise, the present work demonstrates that new fabrication approaches in combination with bio-inspired motion can be used to pave the way for a new set of tools for conducting autonomous vascular surgery, for instance, to specific locations inside the heart [123].

Although the present work focuses on treatment of aneurysms in cerebral blood vessels, the described steerable device can be extended to treat or diagnose a broader set of pathologies through application to other procedures that also require fine, dexterous manipulations. Similar challenges of small vessel diameter and complex vessel tortuosity in addition to vessel occlusions are encountered in interventional cardiology and general vascular surgery. Non-vascular applications include pulmonary nodule biopsy during bronchoscopy in interventional pulmonology, where an analogous branching airway system with progressively smaller branches also involves a navigational procedural challenge. The soft material steerable device presented here has the potential to facilitate a number of procedures that require sub-millimeter diameter catheters in other delicate surgical locations.

## MATERIALS AND METHODS

### Detailed description of the fabrication process

The process of fabricating the steerable catheter tip is complicated by the challenges of working at this scale and the need to produce a smooth, uniform, and fluid-tight multilayer structure. The basic procedure is illustrated in Fig. 7, starting with a-d) fabrication of the molds, followed by e-f) mold alignment, g-i) formation of the relatively stiff thin polymer films present on the inner and outer diameter of the steerable tip, j) the alignment of metal rods as casting structures for the hydraulic passages, and finally k) producing the l-m) steerable tip via casting and demolding of the silicone rubber. The capillary tubes and glass strips were bonded to the microscope slide using using an ultraviolet (UV) light cured epoxy (NOA60, Norland Inc., Cranbury, NJ USA). The molding material, a two-part polyurethane plastic, (Smooth-Cast^®^ 327, Smooth-On, Inc., Macungie, PA USA), selected for its hardness and smooth finish, was poured over the assembly and degassed. Upon curing, the polyurethane plastic was de-bonded from the glass tubing to produce one-half of the mold. Two halves were placed facing each other with a custom manufactured glass capillary tube (Vitrocom, Inc, Mountain Lakes, NJ USA) between them for alignment, with a vise to clamp the assembly. The 900 *µ*m OD, 400 *µ*m ID glass capillary tube has four 60 *µ*m channels in the wall for later introduction of wire that serve to mold the fluid passages used to steer the tip. Dragon- Skin^®^ 10 SLOW (Smooth-On, Inc., Macungie, PA USA) was pumped into the mold to fill it. Dry compressed air at *∼* 50 psi was then blown through the mold to clear the mold, leaving a thin coating of Dragon-Skin upon the mold surfaces.

**Figure 7:**
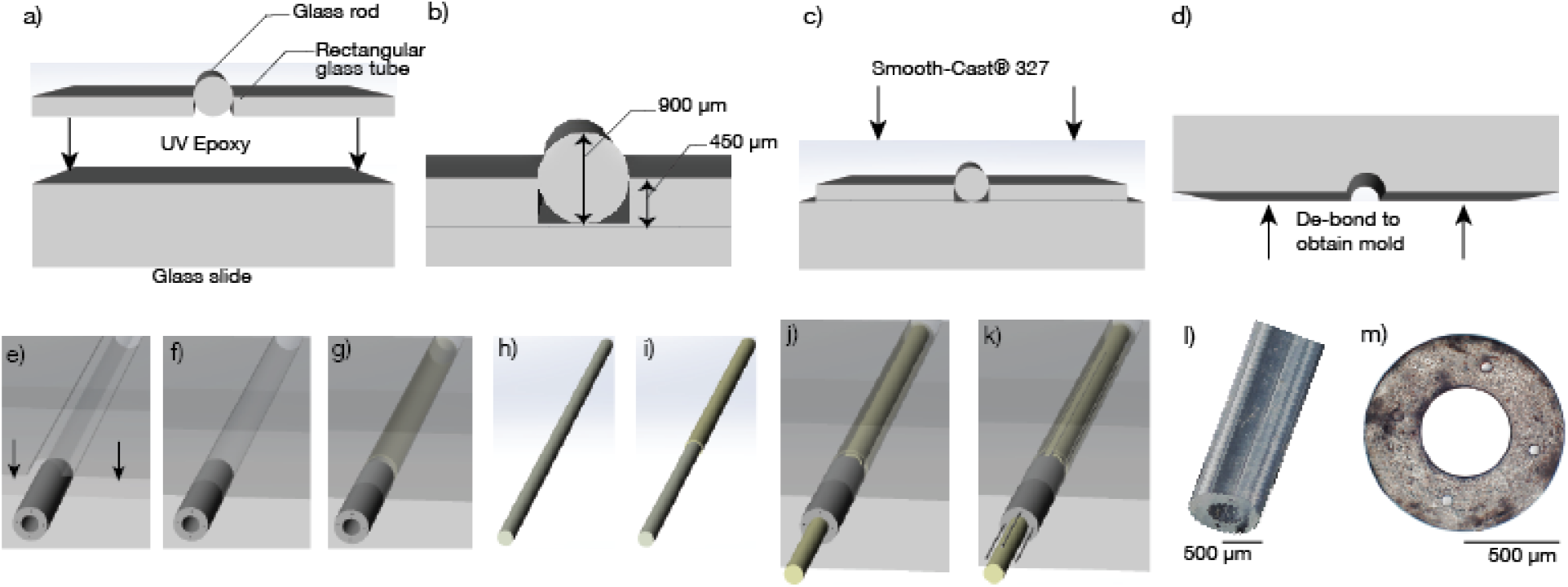
Steerable catheter tip fabrication process. a) Circular glass rods (diameter 900 *µ*m) were placed next to 450 *µ*m-thick rectangular glass sheets and b) bonded to a microscope glass slide using UV epoxy. c) Smooth-Cast^®^ 327 was poured over the assembly and, upon curing, d) de-bonded to obtain one-half of the complete mold. The mold is formed by e,f) clamping two such halves facing each other with a custom glass capillary tube (outer diameter of 900 *µ*m, inner diameter of 400 *µ*m, four 50 *µ*m channels in the wall) to provide alignment. Relatively stiff Dragon-Skin 10 SLOW is first injected into the mold and then blown out with air to produce a thin layer g) on the inside of this mold. A h) 400 *µ*m glass capillary tube is likewise i) spin-coated with the same material and j) introduced into the mold with k) four wires that represent the fluid passages for tip steering. The vacant regions are filled with Dragon-Skin 10 SLOW mixed with Hexane. The result of this process is a l,m) multilayer soft polymer structure.

Separately, a 400 *µ*m diameter glass capillary tube was coated with release agent (Ease Release^®^205 (Smooth-On, Inc., Macungie, PA USA); *see* Fig. 7h-i). The tube was spin coated with Dragon-Skin^®^ 10 SLOW at 2000 rpm for 25 s and left to cure for 24 h. The Dragon-Skin^®^ 10-coated glass-capillary tube was placed in the central lumen of the mold structure. Four 2% rhenium-tungsten rods (Mitaka Co., Saitama, Japan), 50 *µ*m in diameter, chosen for their rigidity and smooth surface finish, were then passed through the capillary channels in the multibore glass capillary tube until they extended beyond the structure by 15 mm. Dragon- Skin^®^ 10 SLOW, mixed with hexane (296090, SigmaAldrich, St. Louis, MO USA) in a 1:1 ratio was degassed and pumped with a syringe through the open end of the mold to fill it. Once cured, the wires and 400 *µ*m diameter glass capillary tube were carefully withdrawn, leaving behind the cavities for the inflatable microchannels and the central lumen. The Smooth-Cast 327 molds easily detach from the silicone rubber to obtain a steerable tip (Fig. 7l) With precisely defined dimensions and micro-hydraulic channels (Fig. 7m).

### The multilayer structure of the polymer in the tip is important to avoid radial expansion

Without concentrically layering the outer and inner surface of the steerable tip with Dragon- Skin^®^ to provide a relatively stiff surface upon the 1:1 Dragon-Skin-hexane combination that forms the majority of the flexible tip material, the steerable tip exhibits significant radial expansion as indicated in Fig. 8. With the relatively soft 1:1 Dragon-Skin-hexane combination alone (“Non-layered”), the radial expansion is at least four times the layered structure of 1:1 Dragon-Skin-hexane inside a thin film of Dragon-Skin. The surface layer is approximately 25 *µ*m thick as seen in Fig 2d).

**Figure 8:**
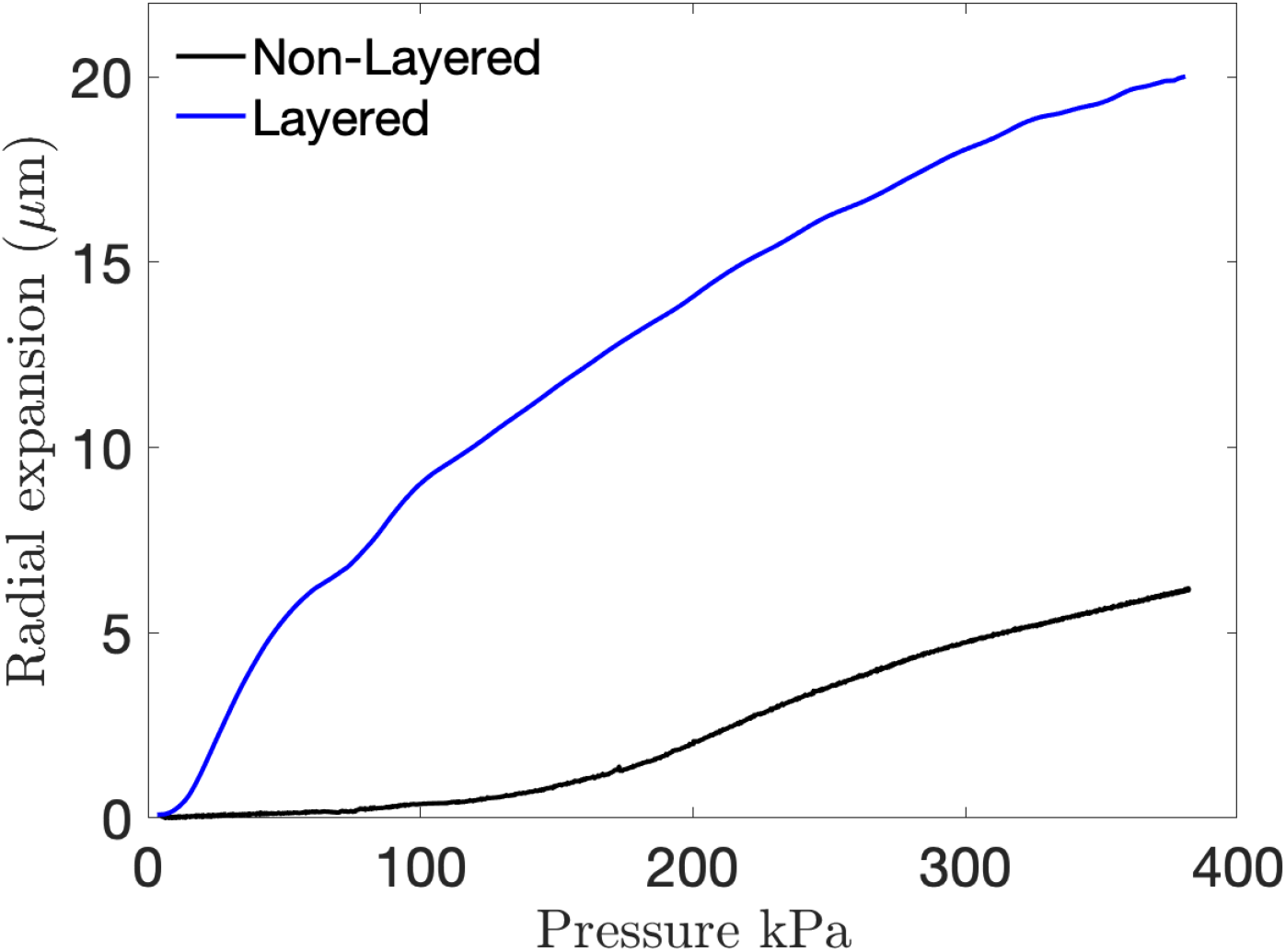
A thin coating of relatively stiff polymer significantly reduces radial expansion. Radial expansion of the steerable tip with and without a relatively stiff 25 *µ*m-thick layer of Dragon-Skin^®^ 10 SLOW upon Dragon-Skin-hexane to form the bulk of the steerable tip structure. The steerable tip fabricated with concentric layers of Dragon-Skin^®^ encompassed by Dragon-Skin^®^ mixed with hexane exhibits significantly smaller radial expansion (5 *µ*m) relative to a steerable tip without the coating (20 *µ*m).

### Assembly of full length devices with transitional stiffness

In each case, the 400 *µ*m diameter central lumen and 50 *µ*m-diameter channels were tethered to a handheld controller through microtubing to luer lock connectors. This process involves connecting polyimide micro-tubing (Microlumen, Oldsmar, FL USA) with the hyperelastic material steerable tip. Pebax^®^ 3533 (Nordson Medical, Huntington Beach, CA USA), selected for its flexibility and durability, was reflowed over the polyimide-Dragon-Skin interface to complete the bonding process. The ratio of polyamide to polyether defines the material’s stiffness or *durometer*. Pebax tube segments of increasing durometer hardness were telescopically connected via standard reflow procedures to obtain full length (160 cm) catheters. The transitional stiffness design was selected to comply with clinical requirements, where lower durometer materials (Pebax 3533, Pebax 4533) are used closer to the cerebral arteries, intermediate durometer materials (Pebax 5033, Pebax 6233) are used in the cardiovascular region, and higher durometer materials (Pebax 7233) are used closer to the insertion region at the femoral artery. While general factors relating to guide catheter selection often come down to personal choice or experience, generally desirable properties in a catheter are a soft atraumatic distal end, a study proximal shaft for support, and multiply graded transitions in rigidity from the proximal to distal ends [81]. In commercially available catheters the lengths and durometer hardness of each section are proprietary. Various systems have been proposed in research [49], some including 12 segments [20], others specifically analyzing respective lengths of stiff and soft sections at the tip [16]. However, due to the subjective preferences, anatomical and clinical case variations, no gold standard exists for lengths of the transitional rigidity zones. Using the representative lengths of the internal carotid artery, common carotid artery, brachiocephalic trunk, descending aorta, abdominal aorta, common iliac, and external iliac artery sections of the catheter were assembled. These were iteratively tested in the ex vivo silicone model and clinician feedback used to modify the lengths to improve “pushability” while maintaining a soft distal end. The final design had Pebax 3533 (10 cm), Pebax 4533 (12 cm), Pebax 5033 (15 cm), Pebax 6233 (23 cm), and Pebax 7233 (100 cm).

### Hydrophillic coating protocol

The assembled micro-catheter is Pebax tubing tethered to a hyperelastic material tip. An internal and external hydrophilic coating (ON-470, Aculon Inc, San Diego, CA USA) was applied to coat the catheter via submerged dip coating. To obtain a clear passageway in the central lumen, compressed air was injected at 2 psi for 2 min after dipping.

### Experimental characterization of the steerable tip

The performance of the steerable tip was quantified through measurements of the tip curvature versus input pressure over time. Combining imagery (FASTCAM Mini AX200, Photron, San Diego, CA USA) with a high precision microfluidics system (OB-1, Elveflow^®^, Paris, France), and custom image post-processing (MATLAB R2020a, MathWorks, Natick, MA, USA), the curvature of the tip was obtained as a function of the input pressure as shown in Fig. 3. The experiment was designed such that the Photron camera triggers recording in synchrony with the input pressure from the Elveflow system. The imposed pressure and camera frame rate were set to be equal at 50 Hz.

### One-to-one motion of the catheter

During surgical intervention, tip deflection requires timely response to the hand motions of the interventionist. The response time, *τ*, can be analytically estimated as *P =* 1*−*exp *<u>^−t^</u>*[79]. The hydraulic channel is a closed system between the input and the hyperelastic material tip. The transient response of the tip deflection to step changes in input pressure in Fig. S1 shows *τ <* 1s, despite 1.6 m of connecting 50 *µ*m diameter microtubing between the two.

### Computational simulations

A finite-element based model was devised (ANSYS, Inc., Canonsburg, PA, USA) to represent the geometrically nonlinear finite deformation of the hyperelastic, nonlinear stiffening material in the catheter tip to produce the results plotted in Fig. S2. Due to the relatively low strains (*<*10%) induced in this application, a two-parameter Mooney-Rivlin model [75, 96] was used to model the material properties. Hyperelastic constants *C*_1_ and *C*_2_ necessary for the model were separately extracted by conducting biaxial membrane tests on thin films [34]. A manual element control with a reduced brick integration scheme and a linear tetrahedron mesh with adaptive sizing was utilized. Boundary conditions include a fixed face at the beginning of the steerable tip and a normal pressure applied within the hydraulic channel. To account for the changes in size and aspect ratio of the elements during the finite deformation of the tip structure, a nonlinear adaptive deformation (NLAD) scheme [6] was used to ensure convergence with the hyperelastic material.

### A representative *ex vivo* model of the vasculature with realistic cardiac flow

Anonymized CT-angiogram DICOM images from a patient with representative vasculature tortuosity for the mean age group treated for intracranial aneurysms were segmented using 3DSlicer [59] to obtain the vasculature in a format suitable for 3D printing. A model of the vascular structure was 3D printed with ABS (Acrylonitrile Butadiene Styrene) plastic. The surface of the 3D printed models was treated with an acetone vapor bath at 40*^◦^*C for 30 minutes to obtain a smooth surface finish. The smoothed 3D model was dip-coated with eight layers of a translucent hyperelastic polymer (SORTA-clear^®^ 40, Smooth-On, Inc., Macungie, PA USA), allowing 24 hours cure time for each layer. The polymer coated model was placed in an acetone bath to dissolve the ABS, leaving behind the *ex vivo* silicone model. The walls of the silicone model were rendered hydrophilic with a surface coating (ON–470, Aculon Inc, San Diego, CA USA) to reduce the friction between the silicone model and the outer walls of the catheter and mimic the lubricity of endothelial cells in the human vasculature.

The use of anonymized DICOM-formatted imagery to produce the vascular in-silicone model was conducted under University of California San Diego’s Institutional Review Board’s approved protocol 171538, entitled “Prospective and retrospective observational study in human subjects undergoing radiology examinations for clinical care”, most recently approved on 28 September 2019.

### Pulsatile flow using a blood flow analog

The *ex vivo* silicone model was connected to an external flow system (FlowTek 125, UnitedBiologics, Santa Ana, CA USA) to generate pulsatile flow and pump a physician verified blood analog (SLIP solution, UnitedBiologics, Santa Ana, CA, USA) to provide a representative validation model to mimic blood pumped by the heart. The pulsatile flow pump enables control of the pulse rate and flow rate. An immersion heater was used to maintain the blood flow analog at a temperature of 37^°^C (98.6^°^C) representative of the clinical environment. Since the silicone model is an isolated segmentation of the endovascular coiling arterial route, the flow rate through the aneurysm location was calibrated to achieve 50 ml/min prescribed by standard measurements of flow rates through cerebral arteries [125]. The temperature of the blood analog was maintained at 37^°^C (98.6^°^F) using an immersion heater with thermostat to represent normal temperature of blood in vessels.

### Flow rate calibration

The pressure at the inlet of the internal carotid artery and exit of the posterior communicating artery (PCOM) (as shown in Fig. S3) was measured using a high resolution pressure sensor (2SMPP-03, OMRON, Kyoto, Japan) [33] at different flow rates generated by the pulsatile flow pump. The pressure difference was used to compute the flow rate assuming laminar flow and therefore the validity of the Hagen-Poiseuille equation to represent the pressure drop-flow rate relationship. The Reynolds number is *∼*50–100, justifying the laminar flow assumption.

### Emulating neuroendovascular surgery, testing, and coil deployment in the *ex vivo* model

Using a female luer connector at the right femoral artery of the silicone model, a guide catheter (ENVOY^®^ XB 6F, Codman & Shurtleff Inc, Raynham, MA USA) was inserted into the silicone model, with its tip parked at the middle of the internal carotid artery (ICA). A rotating hemostatic valve (RHV) was connected to the Envoy hub and the steerable micro-catheter was advanced until the distal end of the Envoy. A second RHV was connected to the hub of the steerable catheter. Both the RHV’s were connected to standard saline at a pressure of 300 mmHg to set up the flush lines in line with standard endovascular neurosurgical protocols [57]. The second RHV was used as the insertion point for the microguidewire (Synchro^2^™, Stryker Neurovascular, Kalamazoo, MI USA). Upon reaching the distal end, both the microguidewire and steerable micro-catheter were advanced in tandem, with the guidewire leading the way to the aneurysm. At the neck of the aneurysm location, the guidewire was retrieved and hydraulic actuation of the steerable catheter used to engage active steering and place the distal end of the steerable tip into the dome of the aneurysm. Once tip was positioned in the geometrical center of the aneurysm, the hydraulic channel was locked using a one way stopcock to hold the steerable tip in a fixed position. Detachable coils (FC-6-20- 3D, AXIUM™ PRIME detachable coils (frame), Medtronic, Irvine, CA USA) were introduced through the steerable micro-catheter’s hub and deployed into the dome of the aneurysm.

### In vivo testing

A 6-F sheath was placed in the left femoral artery using the modified Seldinger technique [102]. Under roadmap fluoroscopic guidance (Siemens Artis Zeego, PA, USA), a 5-F guide catheter (Guider Softip, Boston Scientific, MA, USA) was placed in the right common carotid artery (CCA) over a 0.035 in wire (Glidewire, Terumo, CA, USA). A straight 0.014 in microguidewire (Synchro^2^ ™, Stryker Neurovascular, Kalamazoo, MI, USA) was inserted into the steerable micro-catheter. The micro-guidewire and micro-catheter were advanced to the distal end of the guide catheter in tandem. The 6-F sheath, 5-F guide catheter, and steerable micro-catheter were flushed with continuously running saline using standard rotating hemostatic valves (Touhy-Borst Y-connector). The micro-guidewire was retrieved, the hydraulic steering engaged, and the steerable micro-catheter was advanced to perform *guidewire free* navigation to access the ascending pharyngeal artery. After advancing the steerable portion of the micro-catheter 25 mm into the ascending pharyngeal artery, the pressure in the hydraulic channel was gradually released and the tip returned to its native curvature. The micro-catheter was advanced beyond the radial center of the parotid artery, and then the relevant hydraulic channel was engaged to access to the parotid artery. The catheter was locked in this curvature and position and platinum coils (FC-6-20-3D, AXIUM™ PRIME) were introduced.

## Data Availability

All data needed to evaluate the conclusions in the paper are present in the paper or Supplementary Materials. All data associated with the work will be made available via a UC San Diego Library-maintained link as provided.

https://doi.org/10.6075/J0R49QN8

## Supplementary Materials

Movie S1. 1 Demonstration of catheter steering. Movie S2. 2 *Ex vivo* access, steering, and coil deployment in PCOM aneurysm. Movie S3. 3 *In vivo* porcine model, access, steering, and coil deployment in the parotid artery Movie S4. 4 Access and steering in maxillary artery.

## Acknowledgments

The authors would like to thank Scott Van Voorhis (David Schnurr Associates), Zeus Industrial Products and Aculon, Inc. for their support. The authors would also like to thank the staff at the UC San Diego Center for Future Surgery, and Tammy Livingston (RT). JF wishes to dedicate this paper to the memory of his grandmother, Dorothy Friend, who passed away in large part due to a cerebral aneurysm brought on by a medical misadventure.

## Funding

TCG is grateful to the American Heart Association for grant 16PRE31430005, and the American-Australian Association Sir Keith Murdoch Scholarship in support of this project. The authors are grateful for funding provided by the American Heart Association’s Innovative Project Award (19IPLOI34760705), the state of California’s AB2664 Medical Entrepreneurship Education and Training Grant scheme, UCSD’s Galvanizing Engineering in Medicine program, and the National Institutes of Health via the University of California Center for Accelerated Innovation (NIH/ NCATS UCSD CTRI 1UL1TR001442-01).

## Author contributions

JF devised the initial concept with BY, designed the research, method, testing, and characterization techniques. TG designed and fabricated the device, devised the fabrication methods, and conducted image post processing, characterization, and analysis. TG and JW developed the the representative *ex vivo* silicone model and flow set-up. *Ex vivo* testing and feedback were provided by BY, DSD, AK and AN. The *in vivo* porcine test was conducted by TG, JW, DSD, JSP, and AK. TG, JHW, DSD and JF wrote the paper.

## Competing interests

TG and JF are authors on a patent application US20190209811A1, “Hydraulically driven surgical apparatus,” submitted by the University of California and which covers design and manufacturing methods for the steerable catheter and control system [28].

## Data and materials availability

All data needed to evaluate the conclusions in the paper are present in the paper or Supplementary Materials. Any raw data, images, or video associated with this work may be obtained through reasonable request to the corresponding author.

## Notes

### Author Declarations

The animal (porcine) study was approved by the University of California San Diego Animal Care and Use Committee (IACUC) approved protocols S12204 (from 8/17/2016) and S13289 (from 3/15/2017). The use of anonymized DICOM-formatted imagery to produce the vascular in-silicone model was conducted under University of California San Diego's Institutional Review Board's approved protocol 171538, entitled "Prospective and retrospective observational study in human subjects undergoing radiology examinations for clinical care", most recently approved on 28 September 2019.

