## Supplementary Information for "Soft robotic steerable micro-catheter for the endovascular treatment of cerebral disorders"

### One-to-one motion of the catheter

During surgical intervention, tip deflection requires timely response to the hand motions of the interventionist. The response time, $\tau$ can be analytically estimated as $P = 1 - \exp\frac{-t}{\tau}$. The hydraulic channel is a closed system between the input and the hyperelastic material tip. The transient response of the tip deflection to step changes in input pressure shows $\tau < 1$s, despite 1.6 m of connecting 50 $\mu$m diameter microtubing between the two. Figure S1 shows the deflection of the steerable tip with respect to the input pressure. The input pressure $P$ was scaled by the maximum pressure $P_{max}$ and the bending angle $\theta$ by the maximum bending $\theta_{max}$.

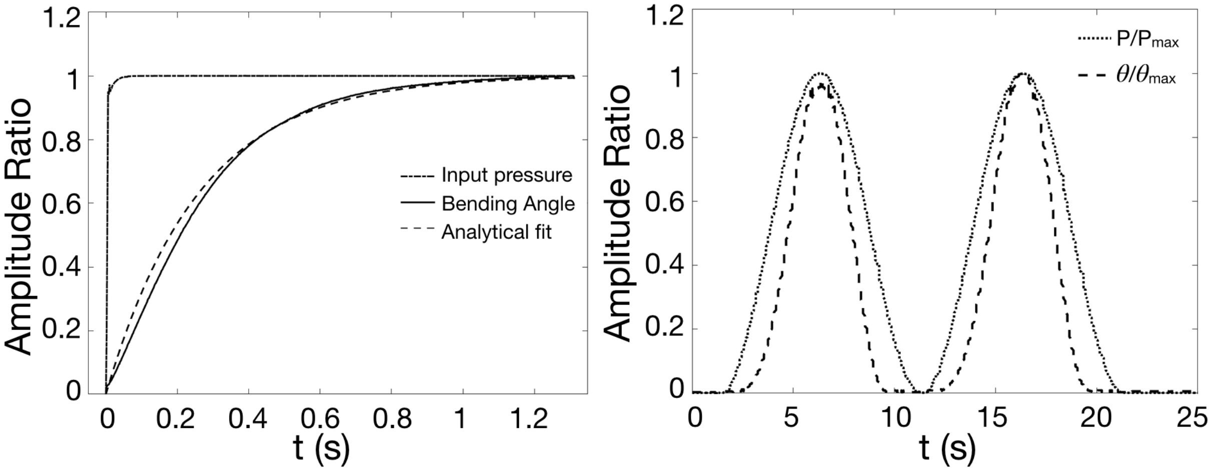


Figure S1: **The distal catheter tip's deflection in response to a commanded input pressure at the proximal end of the 1.6 m-long catheter.** When a step increase in the pressure is imposed at the proximal end of the catheter, the distal tip deflects to 95% of its final response within 1 s to achieve a bending angle of 180°. When a sinusoidal input pressure is gradually imposed, the steerable tip deflects concurrently.

### Computational simulations

A finite-element based model was devised (ANSYS, Inc., Canonsburg, PA, USA) to represent the geometrically nonlinear finite deformation of the hyperelastic, nonlinear stiffening material in the catheter tip. Due to the relatively low strains (<10%) induced in this application, a two-parameter Mooney-Rivlin model was used to model the material properties. Hyperelastic constants $C_1$ and $C_2$ necessary for the model were separately extracted by conducting biaxial membrane tests on thin films. A manual element control with a reduced brick integration scheme and a linear tetrahedron mesh with adaptive sizing was utilized. Boundary conditions include a fixed face at the beginning of the steerable tip and a normal pressure applied within the hydraulic channel. To account for the changes in size and aspect ratio of the elements during the finite deformation of the tip structure, a nonlinear adaptive deformation (NLAD) scheme was used to ensure convergence with the hyperelastic material. The results important to understanding the effect of introducing a pressure into one of the channels is provided in Fig S2.

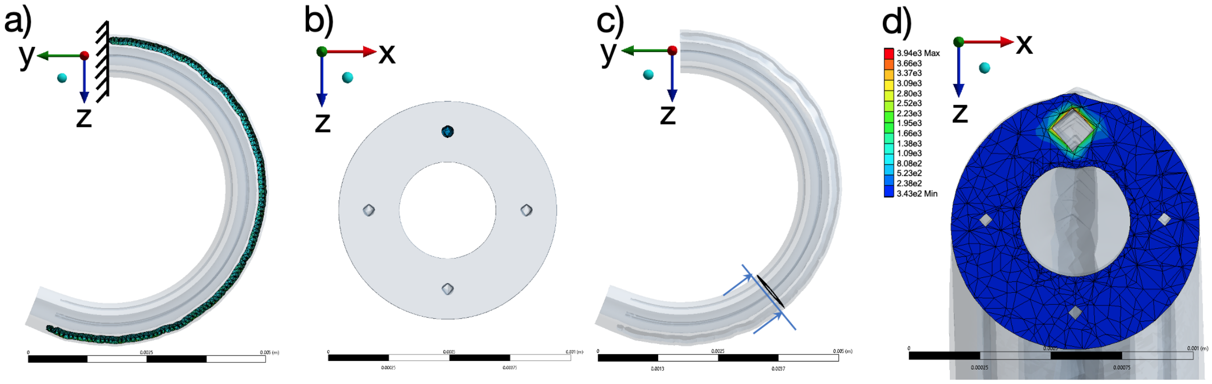


Figure S2: **Computational analysis results of tip deflection.** Result showing the a) pressurized steerable tip with a fixed support and b) cross-section of the tip illustrating the pressurized channel c) illustration of the cross-sectional plane along the length at which the radial pressure is shown in d) achieving an acute at a pressure just under 400 kPa.

### Flow rate calibration

The pressure at the inlet of the internal carotid artery and exit of the posterior communicating artery (PCOM), as shown in Fig. S3, was measured using a high resolution pressure sensor (2SMPP-03, OMRON, Kyoto, Japan) at different flow rates generated by the pulsatile flow pump. The pressure difference was used to compute the flow rate assuming laminar flow and therefore the validity of the Hagen-Poiseuille equation to represent the pressure drop-flow rate relationship. The Reynolds number is ~50–100, justifying the laminar flow assumption.

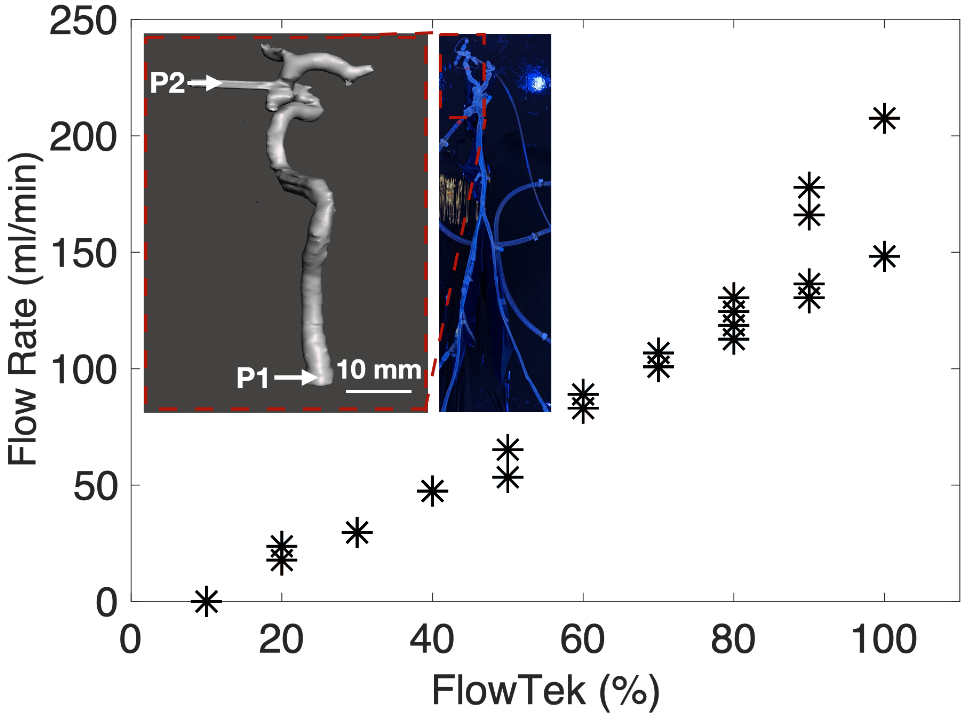


Figure S3: **Calibration of the flow rate through the PCOM aneurysm in the *ex vivo* silicone model.** Inset: illustration of the pressure measurement points in the ex vivo model. Point P1 is at the inlet of the internal carotid artery and point P2 is at the exit of the posterior communicating artery. The pressure drop between these two points, together with the observation that the flow in the interceding artery is laminar, allows us to compute the flow rate via the Hagen-Poiseuille equation. This computed flow rate delivered through the aneurysm depends almost linearly upon the commanded flow rate to the pump.
